# Integration of Mendelian randomisation and systems biology models to identify novel blood-based biomarkers for stroke

**DOI:** 10.1101/2023.03.12.23287170

**Authors:** Tania Islam, Md Rezanur Rahman, Asaduzzaman Khan, Mohammad Ali Moni

## Abstract

Stroke is the second largest cause of mortality in the world. Genome-wide association studies (GWAS) have identified some genetic variants associated with stroke risk, but their putative functional causal genes are unknown. Hence, we aimed to identify putative functional causal gene biomarkers of stroke risk. We used a summary-based Mendelian randomisation (SMR) approach to identify the pleiotropic associations of genetically regulated traits (i.e., gene expression and DNA methylation) with stroke risk. Using SMR approach, we integrated cis-expression quantitative loci (cis-eQTLs) and cis-methylation quantitative loci (cis-mQTLs) data with GWAS summary statistics of stroke. We also utilised heterogeneity in dependent instruments (HEIDI) test to distinguish pleiotropy from linkage from the observed associations identified through SMR analysis. Our integrative SMR analyses and HEIDI test revealed 45 candidate biomarker genes (*FDR* < 0.05; *P_HEIDI_*>0.01) that were pleiotropically or potentially causally associated with stroke risk. Of those candidate biomarker genes, 10 genes (*HTRA1, PMF1, FBN2, C9orf84, COL4A1, BAG4, NEK6, SH2B3, SH3PXD2A, ACAD10*) were differentially expressed in genome-wide blood transcriptomics data from stroke and healthy individuals (*FDR*<0.05). Functional enrichment analysis of the identified candidate biomarker genes revealed gene ontologies and pathways involved in stroke, including “cell aging”, “metal ion binding” and “oxidative damage”. Based on the evidence of genetically regulated expression of genes through SMR and directly measured expression of genes in blood, our integrative analysis suggests ten genes as blood biomarkers of stroke risk. Furthermore, our study provides a better understanding of the influence of DNA methylation on the expression of genes linked to stroke risk.

## 1. Introduction

Stroke is the second largest cause of mortality in the world and one of the major causes of long-term disability [1]. It affects over 15 million individuals worldwide, kills over 5.7 million people, and leaves 5 million people permanently disabled [2]. Stroke is a neurological condition characterised by obstruction of blood circulation in the affected regions of the brain, depriving brain tissues of receiving oxygen and nutrients, which results in the death of brain cells and damage to the central nervous system via cerebral infarction, intracerebral haemorrhage or subarachnoid haemorrhage [3]. Stroke is a heterogeneous disease with multiple subtypes, each with its own aetiology and risk factors [1]. The pathophysiological processes of stroke are very complex, involving numerous molecular events, including energy production failure, excitotoxicity, free-radical mediated toxicity, disruption of blood-brain barriers and immune system dysfunction, which lead to the death of neuronal cells in the brain [4]. The biomarkers for stroke risk assessment of individuals are currently unavailable, warranting the identification of potential biomarkers for stroke risk, which may enhance the stroke risk assessment and develop novel therapeutics.

Past genome-wide association studies (GWAS) have identified several risk loci associated with stroke, including rs2304556 (located in *FMNL2*) and rs1986743 (located in *ARL6IP6*) [5–10]. The latest GWAS meta-analysis of stroke has revealed 89 risk loci associated with stroke [11]. In addition, following GWAS meta-analysis, post-GWAS gene-based association analyses, including MAGMA [12], transcriptome-wide association analysis (TWAS) [13], identified putative genes (e.g., *SH3PXD2A, FURIN)* associated with stroke and stroke subtypes [11]. However, the detection of putative causal genes is very challenging due to complex linkage disequilibrium [14] and the occurrence of the majority of genetic variants associated with complex traits in non-coding regions, which are presumed to affect disease phenotype through gene expression regulation having no direct effect on protein structure or function [15]. Past reports have highlighted that trait-linked genetic variants are more enriched to expression quantitative trait loci (eQTLs) or DNA methylation quantitative loci (mQTLs), suggesting genetic variant-trait association could act through gene expression regulation [16–19].

Several analytical approaches have proposed to integrate the expression quantitative trait loci (eQTLs) data with GWAS, such as transcriptome-wide association study (TWAS) framework and colocalization analysis [20,21], but inferring putative causal variants remains difficult. TWAS framework integrates eQTLs data with GWAS to reveal associations between genes and traits [13]. TWAS studies determine the association of genetically predicted gene expression levels of genes with complex traits; the TWAS framework is unable to provide the effect of causal strength of the variants and horizontal pleiotropy [22]. Colocalisation is also an integrative gene-prioritisation method that integrates eQTLs data with GWAS signals to identify the co-occurrence of variants between pairs of traits [23]. The detected genetic variations (SNPs) are more likely to be functional when GWAS signals colocalise with the eQTL signal [24]. The colocalisation method, however, is not suggested to identify causal associations between pairs of traits.

Summary based Mendelian randomisation (SMR) approach integrates GWAS, expression quantitative trait loci (eQTL), and DNA methylation quantitative trait loci (mQTL) datasets to predict the functional genes which are either pleiotropically (i.e., single genetic variant affecting gene expression and disease phenotype) or potential causally (i.e., single variant affecting gene expression) associated with complex disease [25]. SMR has been used to identify functional genes for many complex diseases and traits, demonstrating SMR as a promising tool for discovering candidate genes linked to complex disorders [25–30].

Multi-omics analyses are well-established to discover functional genes, drug targets, and biomarkers in various complex diseases, including neurological disorders [22,31–34], cancers [35–39], and cardiovascular disease [40]. Multi-omics data integration provides an innovative avenue to bring multi-layer biological information to systematically identify novel insights into the complex pathobiology of diseases [41,42]. However, even with the crucial findings from past GWAS studies, the functional implications and biomarker potentials of the causal genes and loci in stroke risk are largely unknown. Therefore, an approach to systematically integrate post-GWAS analysis with gene expression regulation is desirable for discovering potential biomarkers for stroke risk that may be useful for risk assessment and monitoring. Our bioinformatics approach, which integrates the largest multi-stage multi-omics data integration for stroke, including eQTLs, mQTLs, GWAS, genome-wide gene expression profiling, and functional enrichment analysis, is substantially different than other studies.

Herein, our bioinformatics approach combines SMR method with multi-omics systems biology approaches to identify potential biomarkers, pathways, and therapeutic targets for stroke. We performed an integrative systems biology analysis by combining GWAS summary data for stroke and stroke subtypes with blood cis-eQTL and cis-mQTL data to identify pleiotropic or potential causal genes associated with stroke risk. We have identified putative functional genes and DNA methylation sites associated with stroke risk. Of those candidate functional genes, ten genes were differentially expressed in stroke versus controls, suggesting potential biomarkers of stroke risk assessment. Functional enrichment analysis suggests gene ontologies and pathways involved in stroke, which enhances our understanding of additional biological routes of stroke.

## 2. Materials and Methods

### 2.1 Study Design

**Fig. 1** illustrates the overall analytical approach of this study. To identify pleiotropic or potential causal genes and DNA methylation sites associated with stroke risk, we performed network-based integrative SMR analysis by integrating GWAS summary statistics of stroke from the MEGASTROKE consortium (http://megastroke.org/) with cis-methylation quantitative trait loci (cis-mQTLs) and cis-expression quantitative trait loci (cis-eQTLs) data. Our study included three distinct SMR tests. Firstly, we utilised cis-mQTL data as instrumental variables (IVs), DNA methylation as the exposure, and stroke risk as the outcome to identify the potential pleiotropic or causal association between DNA methylation and stroke risk. Secondly, we used cis-eQTL data as the instrumental variables (IVs), gene expression as the exposure, and stroke as the outcome to determine the potential pleiotropic or causal association of gene expression with stroke risk. Thirdly, we used two molecular traits SMR to assess the putative pleiotropic association between DNA methylation and gene expression, where cis-mQTL acted as the instrumental variables (IV), DNA methylation (mQTL) as exposure, and gene expression (eQTL) as the outcome. **Fig. 2**. represents the potential pathways linking genetic variants to stroke risk.

**Fig. 1.**
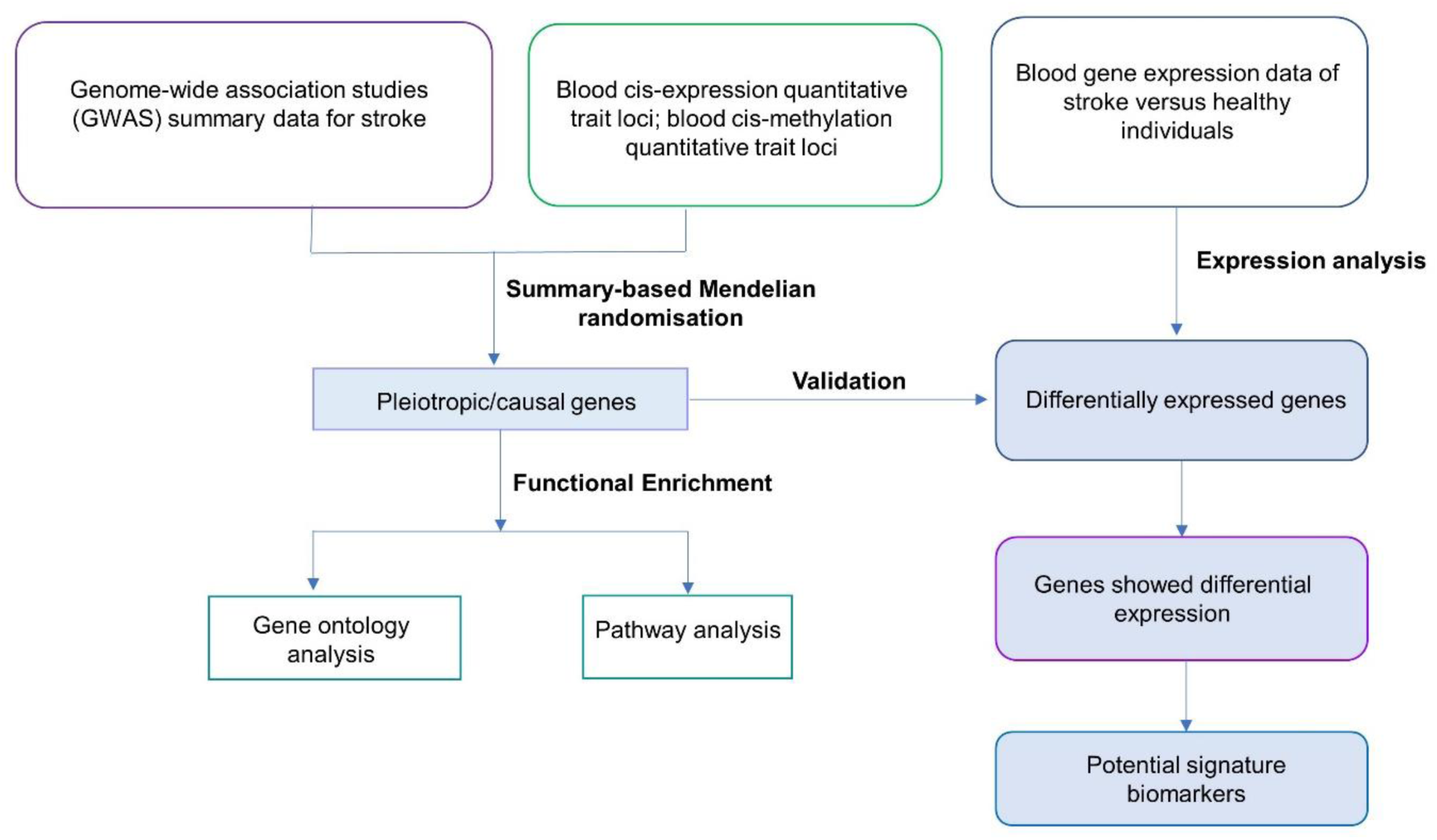
The present study employed a multi-stage analysis methodology. The integrative summary based Mendelian randomisation (SMR) analysis of GWAS summary data of stroke with blood cis-expression quantitative trait loci (cis-eQTLs)/cis-methylation quantitative trait loci (cis-mQTL) data was performed to identify potential causal or pleiotropic gene biomarkers of stroke. Functional enrichment analysis of genes was performed to identify significant gene ontologies and molecular pathways. Differential expression analysis was done on transcriptomic gene expression data of stroke versus healthy individuals. Several SMR- identified genes were differentially expressed in the blood of stroke patients.

**Fig. 2.**
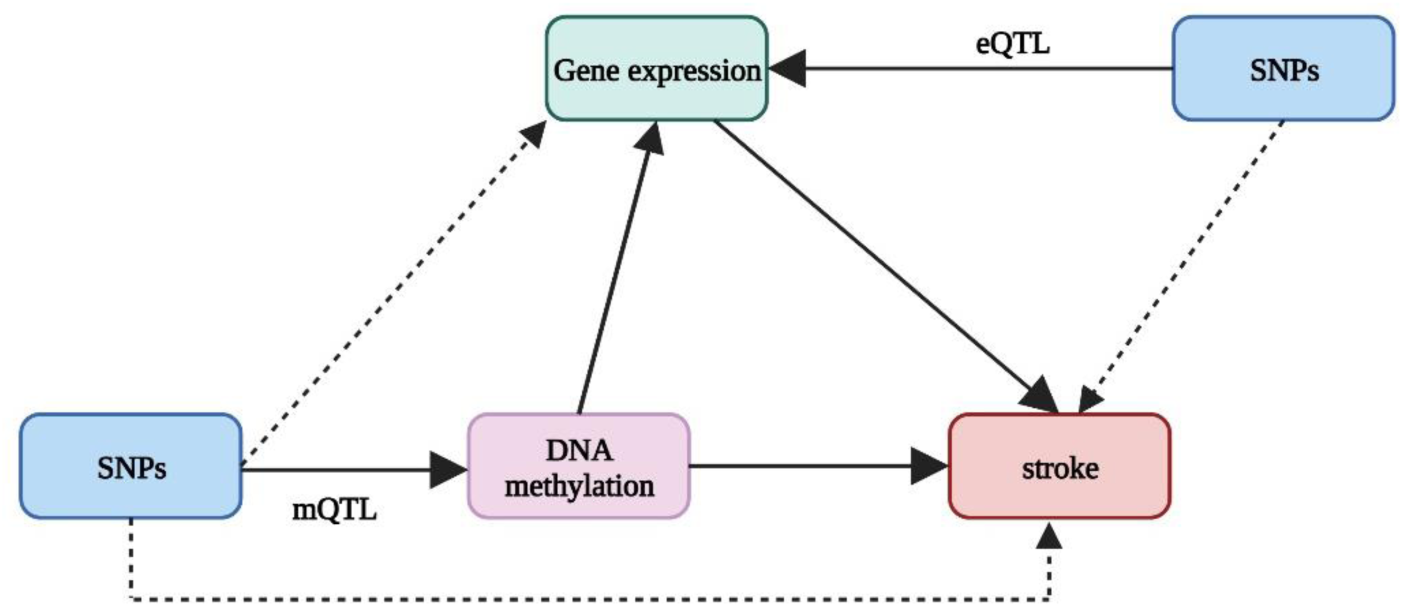
A schematic illustration showing the pleiotropic or causal associations between genetic variants and stroke risk. Firstly, genetic variants can influence stroke risk mediated by DNA methylation. Secondly, genetic variants can influence stroke risk mediated by gene expression. Thirdly, genetic variants can influence stroke risk mediated through genetic regulation of gene expression by DNA methylation. The DNA methylation quantitative loci (mQTL) and expression quantitative loci (eQTL) show genetic variants linked to DNA methylation and gene expression, respectively. The figure was created with BioRender.com.

### 2.2 Data description

Table 1 provides the summary of all the datasets used in this analysis.

**Table 1.**
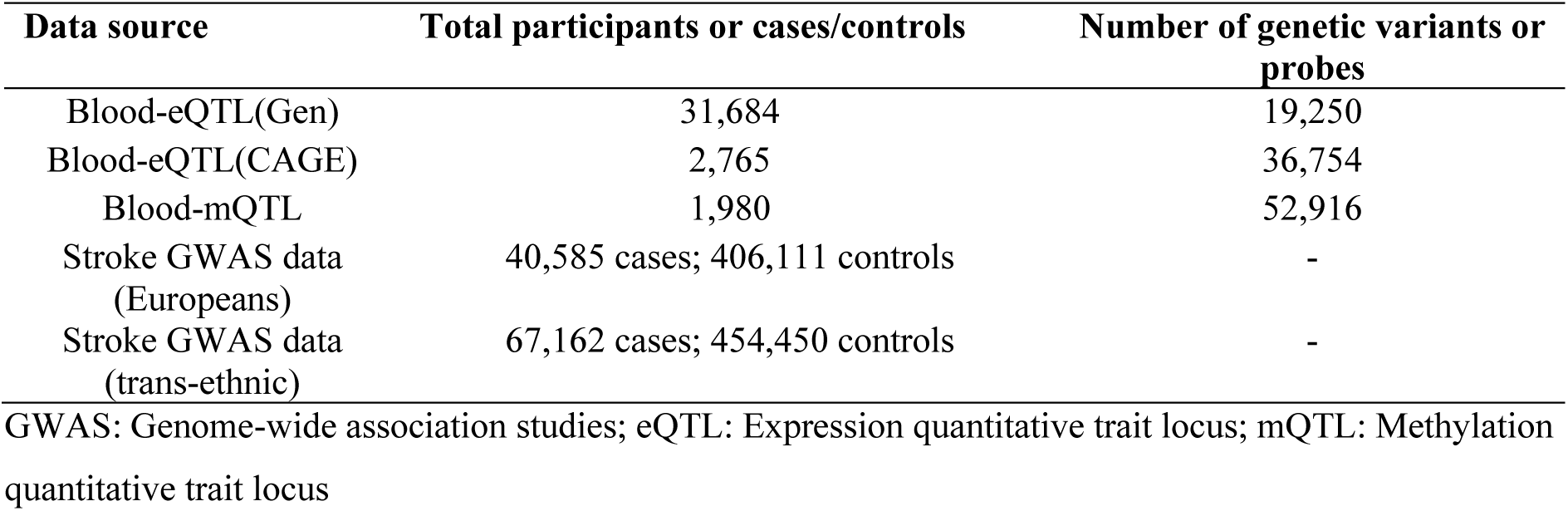
Basic information on the eQTL, mQTL, and stroke GWAS data.

#### 2.2.1 Expression quantitative trait loci data

We used the largest whole blood expression quantitative trait loci (eQTL) dataset from the eQTLGen consortium, which includes cis-eQTLs data for 19250 genes expressed in whole blood obtained from 31,684 individuals. The eQTLs data (i.e., SNP-gene pair) were available for ≥2 cohorts where the SNP-gene distance of ≤1 MB was tested as described previously [43,44]. In addition to this blood eQTL data, we also used another blood eQTL data from the CAGE consortium, which had a total of 2765 participants [45]. The eQTL CAGE dataset used in this study served as the replication of findings obtained from the eQTLGen dataset. The eQTLGen and CAGE eQTL datasets were downloaded on 14 April 2022. The eQTL datasets used in this study are publicly available in the SMR software data resource section (https://yanglab.westlake.edu.cn/software/smr/#eQTLsummarydata).

#### 2.2.2 DNA methylation quantitative trait loci data

DNA methylation quantitative loci (mQTL) are genetic variants that affect DNA methylation levels of a particular transcript. We obtained mQTL summary data of blood from a meta-analysis by McRae et al of the Brisbane Systems Genetics Study cohorts with 614 individuals [46] and Lothian Birth Cohort with 1366 individuals [47,48]. The mQTL data were limited to DNA methylation probes with ≥1 cis-mQTL with at least a cis-mQTL at P < 5e-8 and only SNPs within 2 Mb distance from each probe were available. The mQTL data was downloaded on 14 April <u>2022</u> from https://yanglab.westlake.edu.cn/software/smr/#mQTLsummarydata.

#### 2.2.3 Stroke GWAS summary statistics data

We downloaded GWAS summary statistics for stroke and stroke subtypes from a meta-analysis of GWAS data by Malik et al. [49] released by the MEGASTROKE consortium (http://megastroke.org/). The details of the study have been described previously[49]. Briefly, a fixed-effects meta-analysis was done for GWAS from only European participants consisting of 40,585 cases and 406,111 controls. Then, a fixed-effects trans-ethnic meta-analysis was done including all samples (67,162 stroke cases and 454,450 controls) which consisted of participants of European (40,585 cases; 406,111 controls), East Asian (17,369 cases; 28,195 controls), African (5,541 cases; 15,154 controls), South Asian (2,437 cases; 6,707 controls), mixed Asian (365 cases; 333 controls), and Latin American (865 cases; 692 controls) ancestry [49]. The results of any stroke, as well as the subtypes of stroke, were reported in distinct files which consist of any stroke (AS) cases (67,162), any ischemic stroke (AIS) cases (60,341), and large artery stroke (LAS) cases (6,688), cardioembolic stroke (CES) cases (9,006), and small vessel stroke (SVS) cases (11,710). The datasets were downloaded on 14 April 2022.

### 2.3 Summary-based Mendelian randomisation and heterogeneity in dependent instruments analysis

We used the SMR method described by Zhu et al. [25] and details of the methods described in the original publication by Zhu and co-workers [25]. Briefly, SMR is a data-driven approach used to determine the associations between genetically regulated traits- gene expression or DNA methylation and outcomes of interest- disease phenotypes using genetic variants such as single nucleotide polymorphisms (SNPs) as instrumental variables (IV) [25]. In principle, SMR uses the SNPs association statistics linked to disease phenotype (outcome) regressed on SNPs association statistics linked to gene expression (exposure) to estimate the effect of an increase in gene expression of a particular transcript on the disease phenotype. So, let y be the outcome (e.g., disease phenotype), x be the exposure (i.e., gene expression or DNA methylation), and z be the instrumental variable (i.e., SNPs). SMR determines the effect of gene expression or DNA methylation (exposure) on disease phenotype (outcome) as the ratio of the calculated effect of instrumental variables (SNPs) on disease phenotype (b_zy_) and calculated effect of instrumental variable (SNPs) on gene expression (b_zx_), which can be expressed as (b_xy_ = b_zy_/b_zx_) with no non-genetic confounders [25]. SMR predicts the putative functional genes either pleiotropically (i.e., single genetic variant has effect on both gene expression and disease phenotype), potential causally (i.e., single variant has direct effect on disease phenotype mediated by gene expression) or linkage (two shared genetic variants where one variant has effect on gene expression and another variant has effect on disease phenotypes) [50]. As stated above, an observed association in SMR test could be due to two separate underlying causal variants due to linkage disequilibrium (LD) where one variant has effect on gene expression and another variant has effect on phenotype. This scenario is called linkage. To identify an actual underlying single causal/pleiotropic genetic variant/SNPs from linkage in SMR analysis, we applied a heterogeneity in dependent instruments (HEIDI) test to distinguish the linkage from pleiotropy in the observed association [25]. It was implemented against the null hypothesis that there is a single causal variant deriving the association between exposure (i.e., gene expression/DNA methylation) and outcome (disease phenotype). We excluded the SMR association results when HEIDI test determined the significant heterogeneity of top cis-eQTLs in LD with two distinct causal variant (*P_HEIDI_* <0.01), suggesting the presence of linkage in the observed associations in SMR analysis [25][51]. Therefore, we used *P_HEIDI_* >0.01 as a cut-off to exclude linkage from pleiotropy [51]. The pleiotropic and causal genes are of biological interest for functional characterisation in follow-up studies to develop mechanistic insights of genes in stroke pathogenesis. We did three SMR analyses including 1) SMR analysis with cis-eQTL and stroke GWAS to identify association of gene expression change of each gene and stroke risk; 2) SMR analysis with cis-mQTL and stroke GWAS to identify association of change of DNA methylation levels and stroke risk; 3) two molecular traits SMR analysis using cis-mQTL (exposure) and cis-eQTL (outcome) to identify association of change of DNA methylation level with gene expression. Therefore, our SMR analyses yielded pleiotropic associations between DNA methylation, gene expression, and stroke risk. We used the default parameters in SMR analysis (e.g., minor allele frequency [MAF] > 0.01, *PeQTL* < 5 × 10^−8^, eliminating SNPs having low LD or not in LD [r2 < 0.05] and high linkage disequilibrium [LD, r2 > 0.9] with the top associated eQTL. We adjusted the raw *p-value* yielded in SMR analyses with Benjamini-Hochberg [52] method implemented in the R environment to control the false discovery rate (*FDR*) for multiple testing error . We employed the SMR method in command line Linux environment (https://cnsgenomics.com/software/smr/). To note, we annotated the CpG sites with the closest genes using Illumina HumanMethylation450 BeadChip (https://www.ncbi.nlm.nih.gov/geo/query/acc.cgi?acc=GPL13534).

### 2.4 mRNA expression levels of SMR-identified genes leveraging blood transcriptomics of stroke cases versus healthy individuals

We have obtained a microarray gene expression dataset (accession no: GSE58294) containing human peripheral whole blood gene expression of cardioembolic stroke and healthy controls from the NCBI gene expression omnibus (NCBI-GEO) database [53].GSE58294 was deposited in NCBI-GEO by Stamova et al. [54] [PMID: 25036109]. GSE58294 dataset contained 23 healthy controls and 23 cardioembolic stroke patients using Affymetrix U133 Plus 2.0 microarray technology [54]. The blood samples were taken at three time points following stroke (n = 23), at 3 hours, 5 hours, and 24 hours. The first blood was drawn before treatment (≤ 3 hours) and consecutive blood samples were drawn following treatment with 5 hours and 24 hours of post-onset of stroke. Herein, we analysed differential gene expression patterns at all three time points. We used GEO2R online software integrated in NCBI-GEO database to identify differential expression of genes in stroke cases versus healthy individuals. The differentially expressed genes which passed adjusted *p-value* <0.05 and absolute log fold change >0.5 as statistically significant.

### 2.5 Functional enrichment analysis of the SMR identified genes

We performed gene set enrichment and pathway analysis using bioinformatics tools called ConsensusPath DB [55], accessed on 5 July 2022, to identify significant gene ontology (GO), including biological processes, molecular functions, and potential pathways associated with stroke. ConcensusPathDB is the largest database for pathways, including Wikipathways, Reactome, BioCarta, and the KEGG pathway. The *p-value* obtained from the hypergeometric test was adjusted for the correction of multiple testing errors in the enrichment analysis. GO terms and pathways that passed the adjusted *p-value*<0.05 were considered significant.

## 3. Results

### 3.1 Expression of pleiotropic genes is modulated by stroke risk loci in Europeans

To test the association of genetically determined gene expression and stroke risk in the European population, we used summary based Mendelian randomisation (SMR) analysis, which integrated stroke GWAS summary statistics data with the largest blood eQTL summary data from eQTLGen consortium data (eQTLGen hereafter) and another blood eQTL data from CAGE consortium (CAGE eQTL hereafter), to identify pleiotropic or potentially causal genes associated with stroke. SMR and heterogeneity in dependent instruments (HEIDI) tests demonstrated pleiotropic association of 8 genes with stroke (any stroke; AS), 7 genes with ischemic stroke (any ischemic stroke; AIS), one gene for small vessel stroke (SVS) (**Table 2**), which passed the SMR (*FDR*<0.05) and HEIDI test (*P_HEIDI_* >0.01). Notably, we have also obtained three genes (*PMF1*, *RERE*, and *OBFC1*) in SMR analysis with CAGE eQTL, where *PMF1* and *RERE* were found in SMR analysis using eQTLGen (**Table 2**). The low number of genes were identified in CAGE eQTL due to relatively smaller size data than eQTLGen.

**Table 2.**
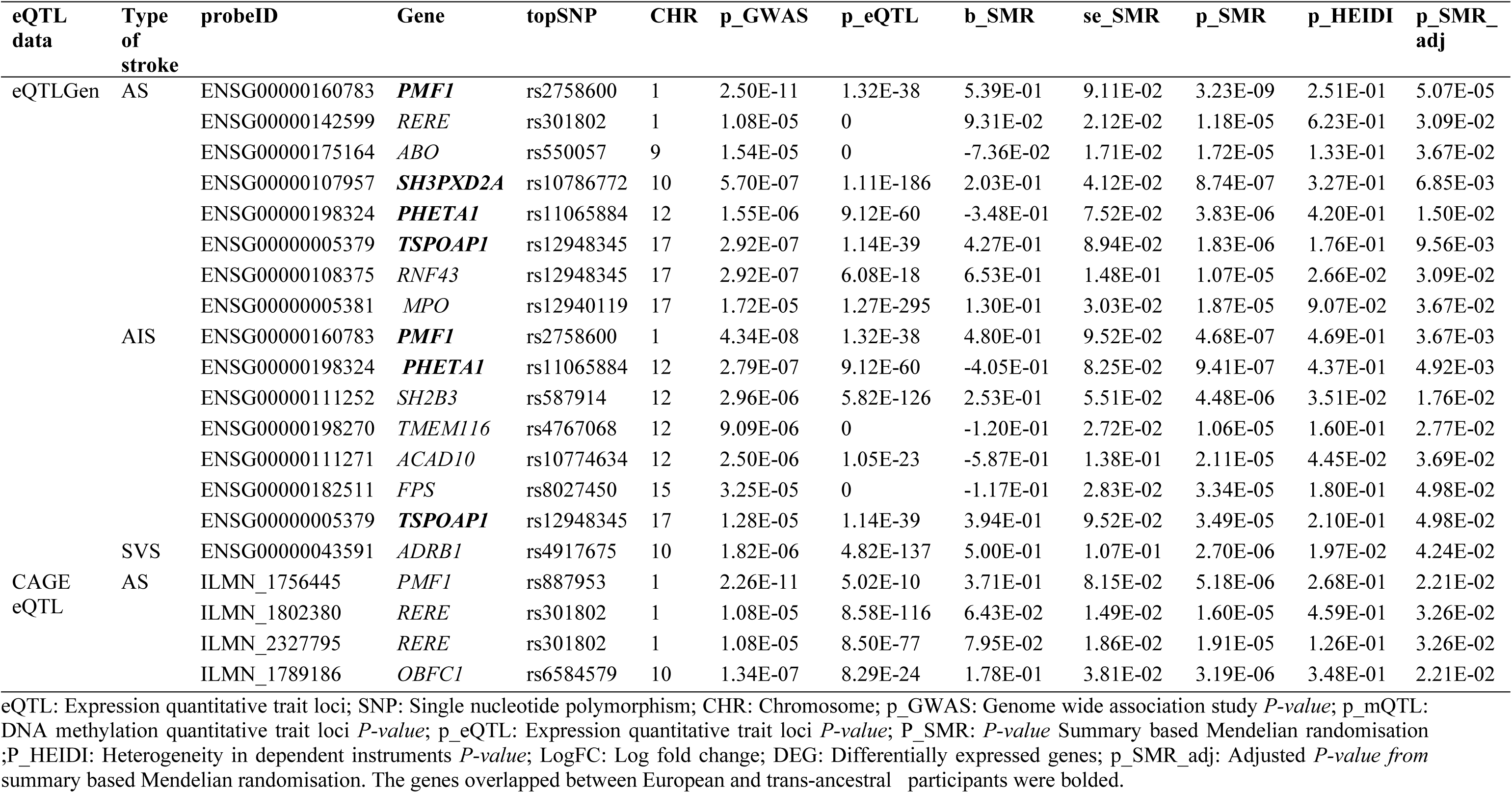
Pleiotropic association between gene expression and stroke risk using blood cis-eQTLs and GWAS of Europeans.

### 3.2 Expression of SMR identified pleiotropic genes is modulated by stroke risk loci in trans-ancestral participants

We used SMR to identify potential genetically determined blood gene expression associated with stroke in trans-ethnic populations and identified several genes with significant pleiotropic or potential causal links with stroke after multiple testing corrections (*FDR* <0.05) and the HEIDI test (*P_HEIDI_*>0.01). Specifically, we found that the expression of 9 genes was pleiotropic or causal for stroke (AS), 5 genes with AIS, and 5 genes with SVS (**Table 3**). Next, in SMR analysis with CAGE eQTL datasets, we found two genes (2 mRNA probes) associated with the stroke while two genes showed potential pleiotropic/potential causal association with AIS. Notably, rs2758600 and rs887953 SNP tagging genes *PMF1* showed significant association with stroke in both SMR analyses using eQTLGen and CAGE eQTLs (**Table 3**).

**Table 3.**
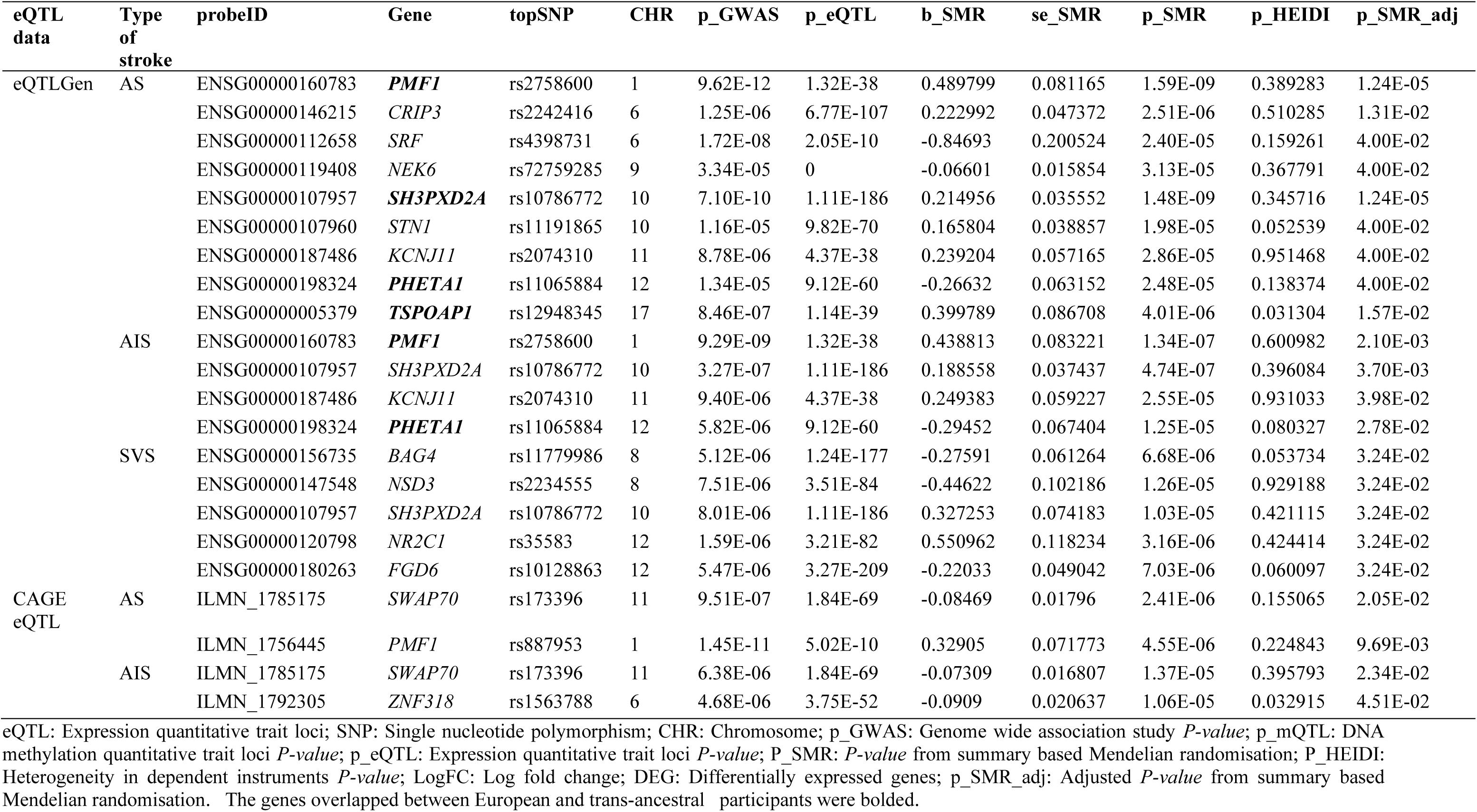
Pleiotropic association between gene expression and stroke risk using blood cis-eQTLs and GWAS of transethnic populations.

### 3.3 Genetically determined expression of common genes overlaps between Europeans and transethnic ancestry

We compared the SMR-identified genes in European and transethnic ancestries to find overlap genes. In SMR with eQTLGen, 4 genes (*PMF1, SH3PXD2A, TSPOAP1,* and *PHETA1*) associated with any stroke were overlapped between European and trans-ethnic populations. In addition, and 2 genes (*PMF1*, *PHETA1*) associated with ischemic stroke were overlapped between European and trans-ethnic populations. In SMR with CAGE eQTL, we found *PMF1* overlapped in both European and trans-ethnic ancestry (Supplementary Table1).

### 3.4 Methylation at CpG sites is modulated by stroke risk loci in European population

To identify CpG methylation sites associated with stroke in European populations, we performed SMR analysis of GWAS summary data for stroke and blood cis-DNA methylation quantitative trait loci (cis-mQTL) data using McRae et al. mQTL summary data [48]. We interrogated the relationship between DNA methylation (i.e., CpG probes) and five different types of strokes. Our analysis identified pleiotropic or potential associations of 22 CpGs with any stroke (AS), 8 CpGs with any ischemic stroke (AIS), and 1 CpG with cardioembolic stroke (CES) (*FDR* <0.05 and *P_HEIDI_>*0.01) (Supplementary Table2). We did not find any significant associations of CpGs with large artery stroke (LAS) and small vessel stroke (SVS) after correction of multiple testing and HEIDI test.

### 3.5 Methylation at CpG sites is modulated by stroke risk loci in trans-ethnic participants

To identify CpG methylation sites associated with stroke in trans-ethnic populations, we conducted SMR analysis of trans-ethnic GWAS summary data of stroke with blood cis-mQTL data as mentioned above. We assessed the association between CpG probes with five types of strokes. We detected 47 CpG methylation sites that showed pleiotropic or potential causal association with AS (Supplementary Table3). In addition, we found that 41CpG methylation sites and 4 CpG methylation sites had significant associations with AIS and CES, respectively, after correction for multiple testing and HEIDI test (*FDR*<0.05 and *P_HEIDI_>*0.01). We did not find any significant CpG sites in LAS and SVS after the correction of multiple tests and HEIDI test (Supplementary Table3).

### 3.6 Multi-omics SMR to prioritise genes mediated by DNA methylation regulating gene expression in stroke

To identify potential associations between DNA methylation and gene expression, we performed two molecular traits SMR analyses by integrating cis-mQTL data with cis-eQTL data. We identified 46,698 associations between 59,975 CpGs and 13,171 mRNA probes after multiple testing corrections (*FDR*<0.05) and the HEIDI test (*P_HEIDI_>*0.01) (Supplementary Table 4). To determine the association between DNA methylation sites that regulate gene expression, we performed an overlapping analysis of CpG methylation sites identified from SMR analysis of mQTL and stroke with the CpG methylation sites identified from SMR analysis of mQTL and eQTL. We found 16 DNA methylated probes regulating the expression of 8 genes (*CAZ1, ACAD10, SLC44A2, SH3PXD2A, PMF1, SUPT4H1, ABO, SLC25A44*) in any stroke (AS), and 5 DNA methylated probes regulating the expression of 4 genes (*ACAD10, SLC44A2, PMF1, SLC25A44*) in any ischemic stroke (AIS) in Europeans (SNP→DNA methylation→Gene expression→Stroke) (Supplementary Table 5).

Next, we also determined the association of DNA methylation regulating gene expression in trans-ethnic ancestry. Similarly, we performed the overlap analysis of CpG methylated sites identified from SMR analysis of mQTL and stroke with the CpG methylated sites identified from SMR analysis of mQTL and eQTL. We identified 34 DNA methylated probes regulating the expression of 16 genes (*CAZ1, SLC38A3, SLC44A2, ILF3, ACAD10, CDKN1A, SH3PXD2A, PMF1, QTRT1, SUPT4H1, HTRA1, SLC25A44, KCNJ11, C9orf84, FBN2, TTBK1*) in AS and 27 DNA methylated probe regulating the expression of 12 genes (*CAZ1, SLC44A2, ILF3, ACAD10, DUS3L, SH3PXD2A, PMF1, QTRT1, SCARF1, SLC25A44, KCNJ11, FAM109A*) in AIS in trans-ethnic ancestry (SNP→DNA methylation→Gene expression→Stroke) (Supplementary Table 6).

### 3.7 Prioritisation of genes associated with stroke risk

Using blood-eQTL and GWAS summary data of stroke, we performed SMR analysis to identify potential pleiotropic or causal associations between gene expression and stroke. Our study found 14 unique gene from Europeans (Table 2) and 15 unique genes from trans-ethnic ancestry (Table 3). Similarly, using blood-mQTL and GWAS summary data of stoke, we also performed SMR analysis to identify pleiotropic or causal association between DNA methylation and stroke. We found total 23 unique CpGs that code for 11 unique genes in European ancestry (Supplementary Table 2) and 58 unique CpGs that code for 24 unique genes in trans-ethnic ancestry (Supplementary Table 3). Considering both gene expression (SMR analysis with eQTL and stroke) and DNA methylation (SMR analysis with mQTL and stroke), we found total 45 unique genes that were pleiotropically or causally associated with stroke risk (Table 4 and Table 5).

**Table 4.** Total unique genes prioritised using summary based Mendelian randomisation.

| Category | European ancestry<br>unique gene | Trans-ethnic ancestry<br>unique gene | Total unique gene<br>between European and<br>trans-ethnic ancestry |
| --- | --- | --- | --- |
| **eQTL prioritised gene** | 14 | 15 | 26 |
| **mQTL prioritized gene** | 11 | 24 | 25 |
| Total unique gene (eQTL+mQTL) between European and trans-ethnic ancestry |  |  | 45 |

eQTL: Expression quantitative trait loci; mQTL: Methylation quantitative trait loci

**Table 5.** Summary statistics of multi-omics prioritised genes associated with stroke risk.

| Gene | topSNP | CHR | A1 | A2 | Freq | b_GWAS | p_GWAS | p_SMR | p_HEIDI | p_SMR_adj |
| --- | --- | --- | --- | --- | --- | --- | --- | --- | --- | --- |
| *PMF1* | rs2758600 | 1 | T | C | 0.343936 | -0.0574 | 4.34E-08 | 4.68E-07 | 0.469249 | 0.003672 |
| *PHETA1* | rs11065884 | 12 | G | A | 0.224652 | -0.0617 | 2.79E-07 | 9.41E-07 | 0.437127 | 0.004919 |
| *SH2B3* | rs587914 | 12 | C | T | 0.202783 | -0.0575 | 2.96E-06 | 4.48E-06 | 0.035064 | 0.017577 |
| *TMEM116* | rs4767068 | 12 | G | A | 0.167992 | -0.0575 | 9.09E-06 | 1.06E-05 | 0.15958 | 0.02767 |
| *ACAD10* | rs10774634 | 12 | G | A | 0.16998 | -0.0615 | 2.50E-06 | 2.11E-05 | 0.044532 | 0.036853 |
| *FPS* | rs8027450 | 15 | T | C | 0.323062 | 0.0454 | 3.25E-05 | 3.34E-05 | 0.179592 | 0.049774 |
| *TSPOAP1* | rs12948345 | 17 | T | C | 0.213718 | 0.0558 | 1.28E-05 | 3.49E-05 | 0.210455 | 0.049774 |
| *RERE* | rs301802 | 1 | T | A | 0.471173 | -0.0413 | 1.08E-05 | 1.18E-05 | 0.623319 | 0.030876 |
| *ABO* | rs550057 | 9 | T | C | 0.282306 | 0.0484 | 1.54E-05 | 1.72E-05 | 0.133303 | 0.036698 |
| *SH3PXD2A* | rs10786772 | 10 | A | G | 0.309145 | -0.0484 | 5.70E-07 | 8.74E-07 | 0.3274 | 0.006852 |
| *RNF43* | rs12948345 | 17 | T | C | 0.213718 | 0.0604 | 2.92E-07 | 1.07E-05 | 0.026604 | 0.030876 |
| *MPO* | rs12940119 | 17 | C | T | 0.172962 | 0.053 | 1.72E-05 | 1.87E-05 | 0.090682 | 0.036698 |
| *ADRB1* | rs4917675 | 10 | C | T | 0.241551 | 0.1247 | 1.82E-06 | 2.70E-06 | 0.019732 | 0.042355 |
| *OBFC1* | rs6584579 | 10 | G | A | 0.362823 | -0.0494 | 1.34E-07 | 3.19E-06 | 0.348092 | 0.022136 |
| *CRIP3* | rs2242416 | 6 | A | G | 0.423459 | 0.04 | 1.25E-06 | 2.51E-06 | 0.510285 | 0.013072 |
| *SRF* | rs4398731 | 6 | A | G | 0.329026 | 0.0469 | 1.72E-08 | 2.40E-05 | 0.159261 | 0.039991 |
| *NEK6* | rs72759285 | 9 | G | A | 0.187873 | 0.0413 | 3.34E-05 | 3.13E-05 | 0.367791 | 0.039991 |
| *STN1* | rs11191865 | 10 | A | G | 0.537773 | 0.0343 | 1.16E-05 | 1.98E-05 | 0.052539 | 0.039991 |
| *BAG4* | rs11779986 | 8 | G | A | 0.258449 | 0.0894 | 5.12E-06 | 6.68E-06 | 0.053734 | 0.03241 |
| *NSD3* | rs2234555 | 8 | A | C | 0.242545 | 0.0838 | 7.51E-06 | 1.26E-05 | 0.929188 | 0.03241 |
| *NR2C1* | rs35583 | 12 | G | A | 0.147117 | 0.1124 | 1.59E-06 | 3.16E-06 | 0.424414 | 0.03241 |
| *FGD6* | rs10128863 | 12 | C | T | 0.148111 | 0.1049 | 5.47E-06 | 7.03E-06 | 0.060097 | 0.03241 |
| *ZNF318* | rs1563788 | 6 | T | C | 0.333002 | 0.0405 | 4.68E-06 | 1.06E-05 | 0.032915 | 0.045126 |
| *SWAP70* | rs173396 | 11 | G | A | 0.405567 | -0.0416 | 9.51E-07 | 2.41E-06 | 0.155065 | 0.020541 |
| *SLC25A44* | rs2842857 | 1 | C | T | 0.351889 | -0.057 | 8.72E-08 | 1.71E-07 | 0.341619 | 0.005076 |
| *CASZ1* | rs880315 | 1 | C | T | 0.356859 | 0.0518 | 9.99E-07 | 2.01E-06 | 0.789051 | 0.023493 |
| *LRCH1* | rs912426 | 13 | C | T | 0.183897 | -0.0629 | 6.22E-07 | 1.01E-06 | 0.028712 | 0.015497 |
| *HTRA1* | rs3793917 | 10 | G | C | 0.195825 | -0.0531 | 2.06E-06 | 3.80E-06 | 0.058166 | 0.019935 |
| *SUPT4H1* | rs2526375 | 17 | G | A | 0.213718 | 0.0597 | 3.62E-07 | 9.37E-06 | 0.213157 | 0.039818 |
| *SLC44A2* | rs2060236 | 19 | G | C | 0.399602 | -0.0465 | 1.54E-06 | 2.15E-06 | 0.631647 | 0.013684 |
| *TMEM51* | rs10927727 | 1 | T | C | 0.161034 | -0.0491 | 6.47E-06 | 7.18E-06 | 0.357166 | 0.023023 |
| *CDKN1A* | rs762624 | 6 | C | A | 0.237575 | -0.0483 | 2.36E-07 | 8.60E-07 | 0.054201 | 0.006601 |
| *ARMS2* | rs11200630 | 10 | C | T | 0.195825 | -0.0453 | 5.35E-06 | 1.87E-05 | 0.301292 | 0.044843 |
| *KCNJ11* | rs5219 | 11 | T | C | 0.352883 | 0.0416 | 1.65E-05 | 2.29E-05 | 0.872022 | 0.049515 |
| *FAM109A* | rs7398833 | 12 | C | T | 0.246521 | -0.0431 | 1.78E-05 | 1.72E-05 | 0.030243 | 0.044531 |
| *SCARF1* | rs28749238 | 17 | G | A | 0.069583 | 0.0808 | 6.25E-07 | 1.38E-05 | 0.13406 | 0.036701 |
| *ILF3* | rs4435370 | 19 | G | A | 0.370775 | -0.0445 | 9.84E-07 | 1.90E-06 | 0.521175 | 0.009819 |
| *DUS3L* | rs45459297 | 19 | A | G | 0.019881 | 0.1283 | 3.13E-06 | 6.39E-06 | 0.033327 | 0.021201 |
| *QTRT1* | rs2229383 | 19 | G | T | 0.400596 | -0.0476 | 4.72E-08 | 1.24E-05 | 0.085246 | 0.033934 |
| *SLC38A3* | rs4688745 | 3 | C | T | 0.441352 | 0.0341 | 1.83E-05 | 2.56E-05 | 0.046776 | 0.044901 |
| *FBN2* | rs73350117 | 5 | C | A | 0.171968 | 0.056 | 5.77E-07 | 1.09E-05 | 0.717204 | 0.025797 |
| *TTBK1* | rs6930689 | 6 | C | T | 0.33002 | 0.0455 | 5.68E-08 | 8.58E-06 | 0.517744 | 0.022191 |
| *C9orf84* | rs823643 | 9 | C | A | 0.439364 | 0.0378 | 6.58E-06 | 1.90E-05 | 0.017133 | 0.034724 |
| *COL4A1* | rs2131938 | 13 | T | C | 0.354871 | 0.0386 | 1.85E-06 | 1.11E-05 | 0.287121 | 0.025797 |
| *PITX2* | rs2739206 | 4 | G | C | 0.302187 | 0.1203 | 1.76E-10 | 1.81E-06 | 0.445129 | 0.028187 |

SNP: Single nucleotide polymorphism; CHR: Chromosome; A1: Effect allele; A2: Other allele; Freq: Frequency of the effect allele; b\_GWAS: effect size from GWAS; p\_GWAS: Genome wide association study *P-value*; p\_SMR: *P-value* from summary based Mendelian randomisation using eQTL/mQTL; p\_HEIDI: *P-value* from Heterogeneity in dependent instruments test; p\_SMR\_adj: Adjusted *P-value* of summary based Mendelian randomisation.

### 3.8 mRNA expression of SMR prioritised gene in the blood of stoke cases versus controls

We directly examined the expression of SMR identified genes in genome-wide blood transcriptomics, leveraging GSE58294 data of stroke versus controls. we identified ten genes (*HTRA1, PMF1, FBN2, C9orf84, COL4A1, BAG4, NEK6, SH2B3, SH3PXD2A, ACAD10*) were differentially expressed between healthy versus stroke individuals at (≤ 3 hours) time point. Of those genes, seven genes (*HTRA1, PMF1, FBN2, C9orf84, COL4A1, BAG4, NEK6,*) were up regulated and *SH2B3, SH3PXD2A,* and *ACAD10* were downregulated in stroke versus controls (*FDR*<0.05; absolute log fold change >0.50) (Table 6) and Fig.3.

**Table 6.** Ten genes from SMR study were differently expressed in stroke cases and control subjects.

| Gene | topSNP | CpG | CHR | P_GWAS | SMR Study |  |  | DEGs(3h) |  | DEGs(5h) |  | DEGs(24h) |  |
| --- | --- | --- | --- | --- | --- | --- | --- | --- | --- | --- | --- | --- | --- |
|  |  |  |  |  | P_mQTL<br>/P_eQTL | P_SMR | P_HEIDI | Log2FC | FDR | Log2FC | FDR | Log2FC | FDR |
| *PMF1* | rs3768276 | cg25465065 | 1 | 1.36E-07 | 0 | 1.37E-07 | 0.014 | 1.038 | 9.23E-10 | 1.084 | 4.78E-12 | 0.926 | 5.08E-10 |
| *FBN2* | rs73350117 | cg23213887 | 5 | 5.77E-07 | 2.11E-20 | 1.09E-05 | 0.717 | 1.028 | 1.23E-05 | 1.122 | 2.05E-06 | 1.384 | 1.56E-07 |
| *C9orf84* | rs823643 | cg01822570 | 9 | 6.58E-06 | 7.03E-43 | 1.90E-05 | 0.017 | 0.925 | 4.34E-03 | 1.215 | 3.62E-06 | 1.630 | 1.80E-06 |
| *COL4A1* | rs2131938 | cg13424422 | 13 | 1.85E-06 | 7.66E-30 | 1.11E-05 | 0.287 | 0.594 | 7.29E-03 | - | - | 0.583 | 2.38E-02 |
| *NEK6* | rs72759285 | - | 9 | 3.34E-05 | 0 | 3.13E-05 | 0.367 | 0.550 | 2.47E-04 | - | - | 0.521 | 1.63E-04 |
| *HTRA1* | rs3793917 | cg25446361 | 10 | 1.25E-06 | 8.39E-96 | 9.04E-03 | 0.383 | 1.355 | 1.39E-04 | 1.511 | 1.15E-05 | 1.930 | 5.50E-07 |
| *BAG4* | rs11779986 | - | 8 | 5.12E-06 | 1.2E-177 | 6.68E-06 | 0.053 | 0.590 | 4.22E-03 | 0.713 | 2.20E-04 | 0.631 | 1.63E-03 |
| *SH2B3* | rs587914 | - | 12 | 2.96E-06 | 5.8E-126 | 4.48E-06 | 0.035 | -0.510 | 2.13E-05 | - | - | - | - |
| *SH3PXD2A* | rs11191833 | cg01727419 | 10 | 1.66E-10 | 0 | 1.77E-10 | 0.396 | -0.588 | 3.89E-04 | -0.710 | 1.04E-06 | -0.639 | 6.29E-06 |
| *ACAD10* | rs642898 | cg08577424 | 12 | 1.37E-07 | 3.3E-187 | 2.17E-07 | 0.039 | -0.837 | 5.76E-05 | -0.502 | 1.61E-02 | -0.524 | 6.20E-03 |
| *SUPT4H1* | rs2526375 | cg14996805 | 17 | 6.68E-07 | 3.99E-19 | 1.42E-05 | 0.026 | - | - | 0.553 | 1.2E-04 | - | - |
| *RERE* | rs301802 | - | 1 | 1.08E-05 | 0 | 3.23E-09 | 0.251 | - | - | 0.679 | 3.59E-04 | 0.644 | 7.52E-04 |

SNP: Single nucleotide polymorphism; CHR: Chromosome; p\_GWAS: Genome wide association study *P-value*; p\_mQTL: DNA methylation quantitative trait loci *P-value*; p\_eQTL: Expression quantitative trait loci *P-value*; P\_SMR: *P-value* from summary based Mendelian randomisation; P\_HEIDI: *P-value* from Heterogeneity in dependent instruments test; DEG: Differentially expressed genes; LogFC: Log fold change; 3h, 5h,24h:Blood samples taken at three time points 3 hours(3h), 5 hours(5h), and 24 hours(24h) following stroke; FDR: False discovery control rate.

**Fig. 3.**
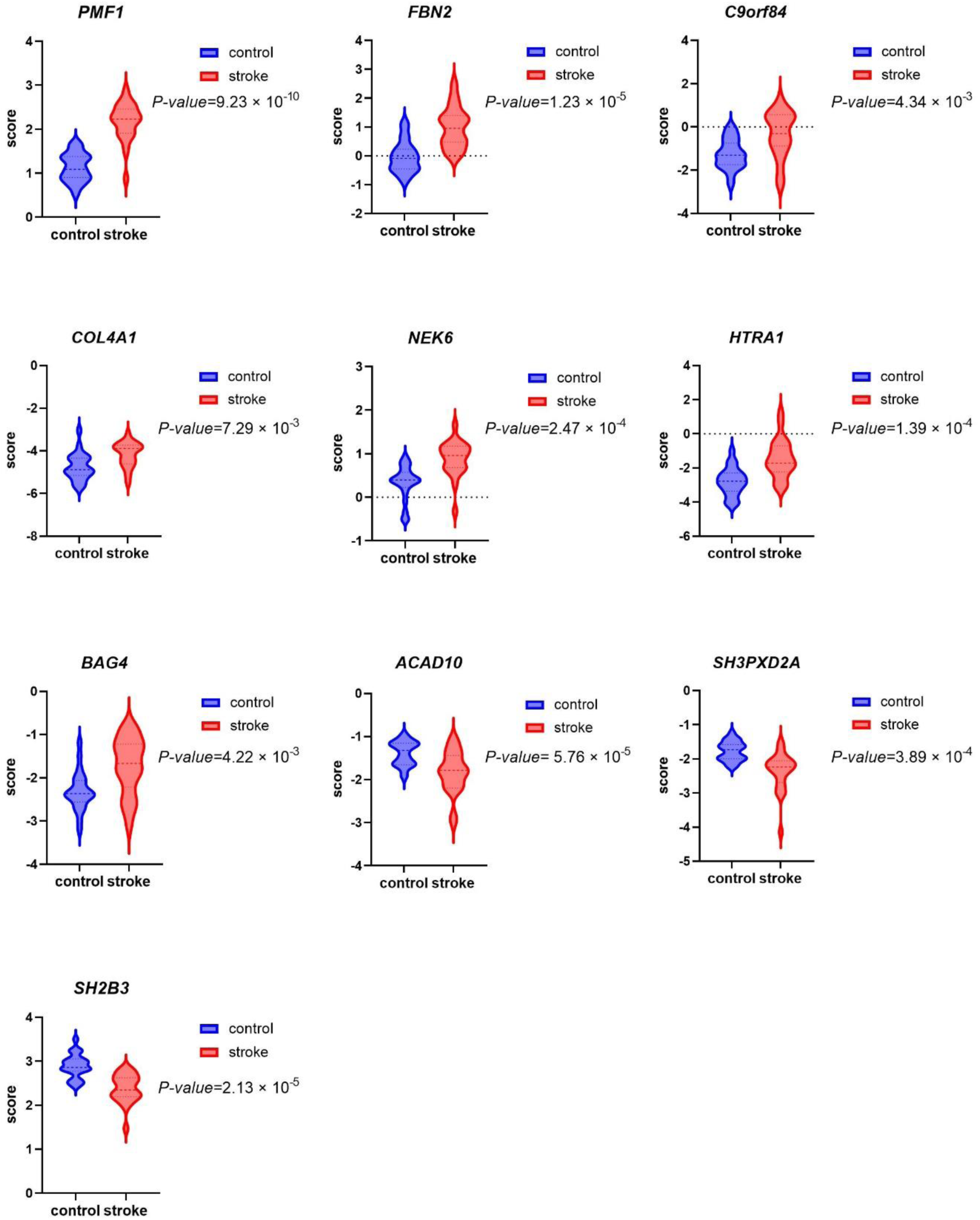
The violin plot represents the significant differential expression of the genes in blood of stroke compared to healthy individuals at ≤3 hours.

Considering gene expression at 5 hours time point, we found nine genes (*HTRA1, PMF1, FBN2, C9orf84, BAG4, SUPT4H1, RERE, SH3PXD2A, ACAD10*) were differentially expressed in stroke where seven genes (*HTRA1, PMF1, FBN2, C9orf84, BAG4, SUPT4H1, RERE)* were up regulated and two genes (*SH3PXD2A, ACAD10*) were down regulated (Table 6). To note, we found seven common genes were differentially expressed in stroke in two time-points (i.e., ≤ 3 hours and 5 hours, respectively). Similarly, we also considered gene expression in 24 hours time point and found ten genes (*HTRA1, PMF1, FBN2, C9orf84, COL4A1, BAG4, NEK6, RERE, SH3PXD2A, ACAD10*) were differentially expressed in stroke where eight genes ((*HTRA1, PMF1, FBN2, C9orf84, COL4A1, BAG4, NEK6, RERE)* were up regulated and two genes (*SH3PXD2A, ACAD10*) were down regulated (Table 6). To note, of those ten genes identified at 24 hours time point, we found nine common genes were also differentially expressed in stroke at ≤ 3 hours time point.

### 3.9 Functional Enrichment

To better understand the potential biological roles of the identified candidate genes in this study, we performed functional enrichment analysis of SMR identified genes. Our functional enrichment analysis of the SMR prioritised genes revealed some significant gene ontology and pathways. They were enriched in some gene ontology terms such as “response to extracellular stimulus”, “cell aging”, “metal ion binding” (**Fig. 4A** and Supplementary Table 7) and pathways, particularly “oxidative damage”, “extracellular matrix organisation”, “transcriptional regulation by the AP-2 (TFAP2) family of transcription factors”, “heart development”, and “coregulation of androgen receptor activity” (**Fig. 4B** and Supplementary Table 8).

**Fig. 4.**
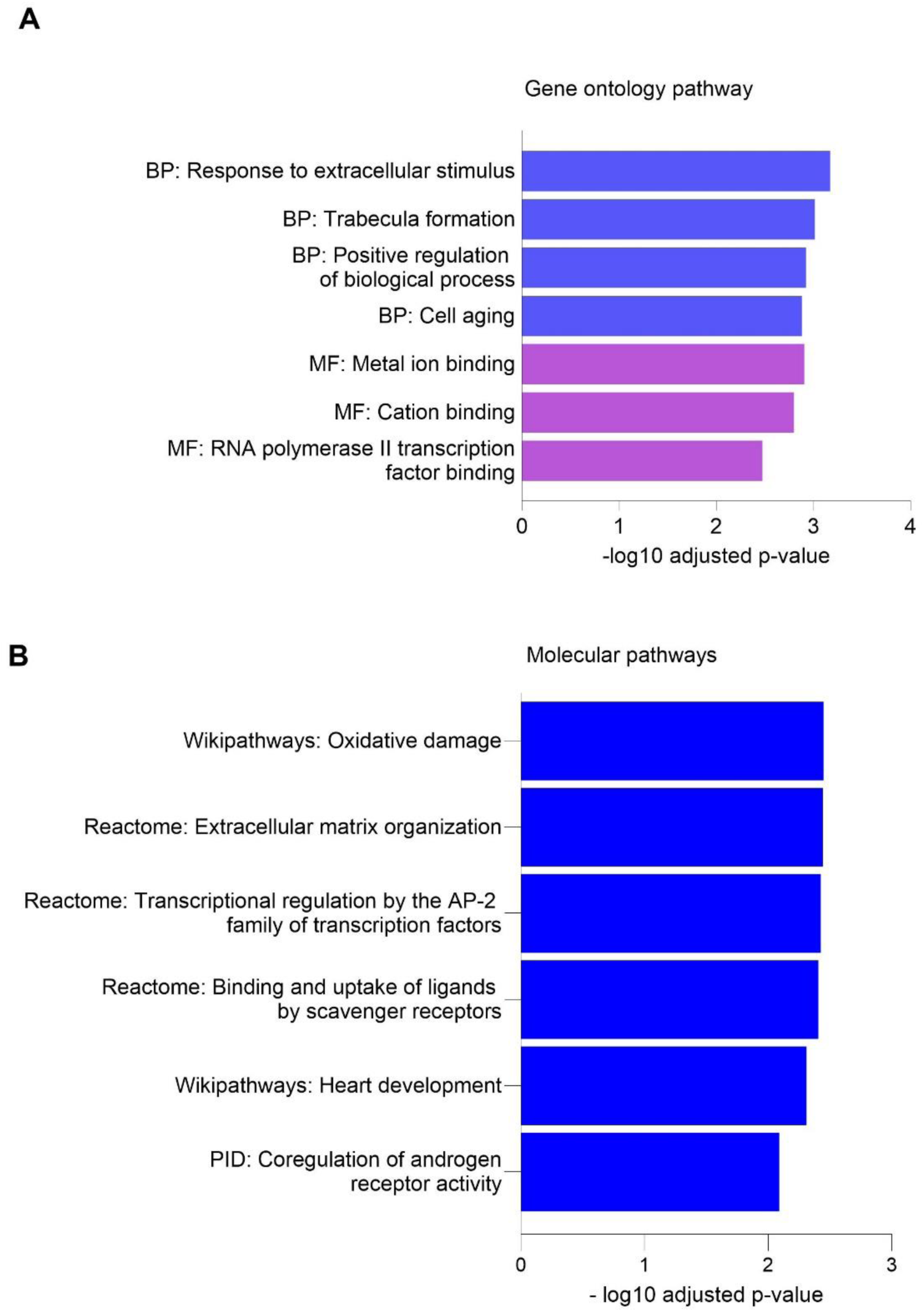
(A) Gene enrichment analysis (blue colour represent biological process and purple colour represents molecular function). (B) The pathway enrichment analysis.

## 3 Discussion

In this present study, we comprehensively integrated multi-omics data, particularly GWAS of stroke, eQTLs, and mQTLs using SMR to identify gene biomarkers of stroke risk. Our network SMR analyses aimed to identify the pleiotropic association of genetically regulated traits- DNA methylation and gene expression with the phenotype of interest. We identified a total of 45 pleiotropic or potential causal gene biomarkers associated with stroke risk. Of those 45 genes, 10 genes (*HTRA1*, *PMF1*, *FBN2*, *C9orf84*, *COL4A1*, *BAG4*, *NEK6, SH2B3*, *SH3PXD2A*, *ACAD10*) were differentially expressed in the blood of stroke patients versus controls, suggesting potential biomarkers of stroke risk. Despite many efforts to identify stroke risk loci, the putative functional gene biomarkers of stroke risk remain unknown. Past studies attempted to explore the risk genes associated with stroke, but the dearth of systems biological approaches undermined the potential to uncover functional risk biomarkers. Therefore, we conducted the largest study to date to systematically search for blood gene biomarkers for stroke risk and validate them in the blood transcriptomics of stroke patients compared to controls. Of those identified putative gene biomarkers, *PMF1, SH3PXD2A*, *SH2B3*, *COL4A1*, and *HTRA1* were previously known as stroke risk loci according to GWAS catalogues, while the causal associations of *FBN2*, *ACAD10*, *C9orf84*, *BAG4*, and *NEK6* were not previously known, suggesting novel gene biomarkers of stroke. The identified genes have been implicated in stroke pathogenesis. *PMF1* is associated with intracerebral haemorrhagic stroke [56,57]. *PMF1* encodes polyamine-modulated factor 1 that plays a significant role in chromosomal alignment and kinetochores formation during mitosis [57]. A previous study found genetic variants of the *SH2B3* gene were significantly associated with the pathogenesis of ischemic stroke, large artery stroke, and coronary artery disease [58]. Past studies reported that mutations in *COL4A1* gene were linked to a wide range of disorders, including haemorrhagic stroke, cerebral small vessel disease, myopathy, glaucoma, and among others [59,60]. Mutation in *HTRA1* is responsible for cerebral autosomal recessive arteriopathy with subcortical infarcts and leukoencephalopathy (CARASIL) and cerebral small vessel disease [61]. CARASIL is a hereditary condition in which small blood vessels in the brain are damaged, resulting in stroke and other disabilities [62]. Moreover, the *HTRA1* gene is also associated with lacunar stroke [63]. *FBN2* gene encodes fibrillin-2 protein, which is involved in tissue elasticity, providing connective tissues with the strength and flexibility to maintain joints and organs in the body [64]. *FBN2* is strongly associated with age-related macular degeneration (AMD) [65], and AMD increases the risk of stroke by 2-fold more than patients without AMD [66]. *SH3PXD2A* (SH3 And PX Domains 2A) is a protein coding gene and was previously associated with ischemic stroke and lacunar stroke [63]. *ACAD10* encodes a component of acyl-CoA dehydrogenase enzymes family, which participate beta-oxidation of fatty acids. The variant of *ACAD10* gene was previously been found associated with ischemic stroke [67], coronary artery disease [68], type 2 diabetes, insulin resistance, and lipid oxidation [69]. The role of *C9orf84* in stroke risk is still unknown. *BAG4* gene encodes for BAG cochaperone 4 proteins which is involved in different biological processes such as cell proliferation, survival, and apoptosis processes. Overexpression of BAG4 protein was observed in different types of cancers [70]. However, the role of *BAG4* gene in stroke risk is still elusive. The risk gene *NEK6* encodes protein kinase which plays a role in apoptosis, cell cycle regulation, chemoresistance, and telomere maintenance [71]. *NEK6* genes promote cell proliferation and develop head neck squamous cell carcinoma [72]. In addition, our study also highlighted that among the ten differentially expressed genes, three genes (*PMF1*, *SH3PXD2A*, *ACAD10*) were modulated by DNA methylation and gene expression in Europeans (SNP→DNA methylation→Gene expression→Stroke) and six genes (*HTRA1*, *PMF1*, *FBN2*, *C9orf84*, *SH3PXD2A*, *ACAD10*) were modulated by DNA methylation and gene expression in trans-ethnic ancestry group (SNP→DNA methylation→Gene expression→Stroke).However, we demonstrated their causal and functional insights as they could be modulated through DNA methylations, which could influence gene expression.

Our study demonstrated the pleiotropic association between gene expression and stroke. Our analysis revealed four genes (*PMF1*, *SH3PXD2A, TPSPOAP1, PHETA1*) were overlapped between Europeans and trans-ethnic ancestry. The trans-ethnic ancestral GWAS meta-analysis consisted of substantial European participants (∼60%) than other ancestries. Next, pleiotropic association between DNA methylation and stroke revealed ten genes (*ARMS2, ACAD10, CASZ1, SH3PXD2A, HTRA1, PMF1, SUPT4H1, SLC25A44, LRCH1, SLC44A2*) were overlapped between European ancestry and trans-ethnic ancestry. To note, we identified *PMF1, HTRA1, ACAD10* and *SH3PXD2A* were differentially expressed in the blood of stroke versus healthy individuals, suggesting potential common biomarkers of stroke risk in both ancestries.

Next, our results revealed important biological insights of the identified genes, including “oxidative damage”, “extracellular matrix organisation”, and “coregulation of androgen receptor activation, suggesting potential mechanisms and pathways involved in stroke pathogenesis [73,74]. It is believed that oxidative stress along with chronic neuroinflammation results in neuronal damage associated with stroke [75]. Oxidative stress develops due to the imbalance between the formation and clearance of free radicals during pathological conditions [76]. It is also linked to the aging processes that increase risk of stroke[77]. The brain cells are more susceptible to oxidative damage due to their excessive metabolic rate and sensitivity to ischemia damage [75,78]. In addition, several pioneering next-generation sequencing studies, particularly transcriptomic and epitranscriptomic studies, have suggested substantial contribution of oxidative damage, neuroinflammation, and angiogenesis regulation pathways in response to physiological alterations in brain arteriovenous malformation, cerebral cavernous malformation, and retinal degeneration [79–82]. The extracellular matrix (ECM) contains collagen, fibrous proteins, elastin, laminins, and microfibrils that give structural support, integrity, and elasticity to the tissue [83]. The structure and expression of ECM alter after a stroke [84,85]. Our study also highlighted some molecular functions, particularly metal ion binding and RNA polymerase II transcription factor binding. Some metal ions are important nutrients for brain function and development [86]. Evidence shows that excessive amounts of some metal ions (e.g., Ca, Zn, Fe, or Cu) contribute to brain injury after stroke. Moreover, numerous exogenous metal ions have also been associated with stroke risk, including cadmium, nickel, arsenic, mercury, and aluminium [86].

There are several strengths in this study. We did the largest integration of multi-omics datasets for stroke to identify candidate biomarkers, including the largest eQTL datasets, mQTL datasets, GWAS datasets, followed by validation of the candidate biomarkers as differentially expressed in blood transcriptomic in stroke compared to healthy individuals [25]. In addition, we validated our candidate biomarkers in real human blood gene expression datasets of stroke versus healthy individuals, suggesting support for blood biomarkers of stroke.

Like other multi-omics approaches, it is important to acknowledge a few limitations as various bioinformatics analyses have been provided in this study, which need to be considered when interpreting the results. We used multiple corrections to limit the false positive rates. As with other SMR investigations, we cannot rule out the potential that any of the associations (crucial SNPs, genes, and DNAm sites) identified in this study were false positives. In addition, we used eQTLs and mQTLs data to investigate the impact of genetic variant on gene expression and DNA methylation using SMR. Although the majority of eQTLs and mQTLs can reveal certain causal variants, the presence of complex linkage disequilibrium structure and secondary signals either from GWAS or eQTLs make it challenging to identify actual causal variant. Although we tried to guard against potential biases in our analyses with the use of cis-eQTL and cis-mQTLs, assessment for heterogeneity (HEIDI test), and multiple error corrections, the actual causal associations could still come from remote variants [28,87,88]. We analysed an independent blood transcriptomic dataset of 23 strokes and 23 controls for the differential expression of potential gene biomarkers. This dataset has a limited sample size. Therefore, we propose future work with a larger sample size for further validation of ten candidate blood biomarkers. There is a lack of publicly available data on global methylation profiling on stroke, therefore, we could not assess whether the DNA methylation level of candidate biomarkers was altered in stroke patients versus healthy controls. Future studies should be conducted to validate these candidates in large-scale human DNA methylation profiling and gene expression profiling datasets to gain a better understanding of the functions of those genes in the aetiology of stroke risk and establish them as biomarkers of stroke before clinical use. We propose further experimental validation of candidate biomarkers using relevant clinical samples in future studies since our approach focused on identifying potential candidate biomarkers for stroke risk.

## 4 Conclusions

Overall, we leveraged large-scale multi-omics datasets using a systems biology approach to identify putative pleiotropic and potential causal gene biomarkers of stroke risk. We identified 45 candidate gene biomarkers by network SMR approach. Of those genes, ten genes were differentially expressed in the blood of stroke patients compared to healthy individuals. Our approach showed the importance of gene expression modulation by DNA methylation in stroke risk. Furthermore, our results suggest potential biological processes and pathways underlying the pathophysiology of stroke. The genetic variants, DNA methylation sites, and genes associated with stroke risk identified in this study demand further wet-lab experimental validation to evaluate their potential utility in stroke assessment.

## Data Availability

All data here analysed are publicly available on SMR software Data resources and MEGASTROKE consortium.

https://yanglab.westlake.edu.cn/software/smr/?fbclid=IwAR2958jo4Akujx3_n15JzcPo5Eh5HwWFRJoQG02ZM9CxNwTnSUSiDRYuE60#SMR&HEIDIanalysis

https://www.megastroke.org/

## Acknowledgments

“The MEGASTROKE project received funding from sources specified at http://www.megastroke.org/acknowledgments.html. The author list of MEGASTROKE is available in Supplementary Table 9

**Conflict of Interest:** “Authors declare no conflict of interest”.

## Supplementary files

Table S1. Significant common genes between Europeans and transethnic ancestry show pleiotropic association between gene expression and stroke.

Table S2. Pleiotropic association between DNA methylation and stroke in the blood of European population.

Table S3. Pleiotropic association between DNA methylation and stroke in blood of Trans-ethnic population.

Table S4. Two molecular traits SMR analysis by integrating cis- DNA methylation quantitative trait loci (mQTLs; exposure) and cis-expression quantitative trait loci (cis-eQTL; outcome).

Table S5. Pleiotropic association between DNA methylation and gene expression and stroke in the blood of Europeans.

Table S6. Pleiotropic association between DNA methylation, gene expression, and stroke in the blood of trans-ethnic ancestry.

Table S7. Significant gene ontology shared between European and transethnic ancestry which were associated with stroke.

Table S8. Significant pathways shared between European and transethnic ancestry were associated with stroke.

Table S9. Members of the MEGASTROKE CONSORTIUM.

